# App-based symptom tracking to optimize SARS-CoV-2 testing strategy using machine learning

**DOI:** 10.1101/2020.09.01.20186049

**Authors:** Leila F. Dantas, Igor T. Peres, Leonardo S. L. Bastos, Janaina F. Marchesi, Guilherme F. G. de Souza, João Gabriel M. Gelli, Fernanda A. Baião, Paula Maçaira, Silvio Hamacher, Fernando A. Bozza

## Abstract

**Background:** Tests are scarce resources, especially in low and middle-income countries, and the optimization of testing programs during a pandemic is critical for the effectiveness of the disease control. Hence, we aim to use the combination of symptoms to build a regression model as a screening tool to identify people and areas with a higher risk of SARS-CoV-2 infection to be prioritized for testing.

**Materials and Methods:** We applied machine learning techniques and provided a visualization of potential regions with high densities of COVID-19 as a risk map. We performed a retrospective analysis of individuals registered in “*Dados do Bem*”, an app-based symptom tracker in use in Brazil.

**Results:** From April 28 to July 16, 2020, 337,435 individuals registered their symptoms through the app. Of these, 49,721 participants were tested for SARS-CoV-2 infection, being 5,888 (11.8%) positive. Among self-reported symptoms, loss of smell (OR[95%CI]: 4.6 [4.4 - 4.9]), fever (2.6 [2.5 - 2.8]), and shortness of breath (2.1 [1.6-2.7]) were associated with SARS-CoV-2 infection. Our final model obtained a competitive performance, with only 7% of false-negative users among the predicted as negatives (NPV = 0.93). From the 287,714 users still not tested, our model estimated that only 34.5% are potentially infected, thus reducing the need for extensive testing of all registered users. The model was incorporated by the “*Dados do Bem*” app aiming to prioritize users for testing. We developed an external validation in the state of Goias and found that of the 465 users selected, 52% tested positive.

**Conclusions:** Our results showed that the combination of symptoms might predict SARS-Cov-2 infection and, therefore, can be used as a tool by decision-makers to refine testing and disease control strategies.

## Introduction

The current COVID-19 pandemic caused by the SARS-CoV-2 requires extensive testing programs to understand the transmission, diagnose, and isolate the positive cases. Given the high mortality and absence of a specific consensual treatment or a reliable vaccine, large testing programs are an essential part of epidemic control. The frequency of testing, however, is very heterogeneous among countries. Brazil currently has the second-highest number of COVID-19 cases, even with lower test rates (59,252 tests per one million inhabitants, as of July 27, 2020)[1].

In a recent paper, Menni and colleagues[2] used information from an app-based symptom tracker from UK and USA and concluded that the combination of symptoms could be used as a screening tool to identify people with a possible positive result for COVID-19. However, little is known about this association and their potential usage as a screening tool in low- and medium-income countries (LMIC) such as Brazil. Thus, our study aims to use the combination of symptoms and machine learning techniques to develop a predictive model that identifies people and areas with a higher risk of SARS-CoV-2 infection. We used data from an app-based symptom tracker known as “*Dados do Bem*”[3], which is an initiative that started available for the state of Rio de Janeiro, one of the centers of the outbreak in the country. Applying our model, we can estimate the proportion of infected participants and then categorize risk levels of infection within the geographical area of the state of Rio de Janeiro, aiming to prioritize tests and optimize the testing program.

## Materials and Methods

### Study design and data source

This study is a retrospective analysis of prospectively collected data from individuals registered in the “*Dados do Bem*” app, which is a large Brazilian initiative that combines an app-based symptom tracker and a public testing initiative for the users. The app interface and the survey questions are provided as supporting information (S1 Fig).

The free smartphone application was launched in Brazil on April 28, 2020. Through a short survey, it collects geo-referenced data from subscribed users, their demographic and occupational characteristics, reported symptoms, as well as whether the participant is a health professional or was in contact with a SARS-CoV-2 infected person. The app then combines the surveyed information and selects individuals for testing. The test used is the antibody WondfoCOVID-19 IgM/IgG test (sensitivity = 86.43%, specificity = 99.57%) [4] and is currently available only for the state of Rio de Janeiro.

### Study population

We included participants registered through the smartphone app from its launch date until July 16, 2020, separating them into two groups: those already selected and tested for the antibody WondfoCOVID-19 IgM/IgG test, and those who responded to the questionnaire but were not tested yet. The tests of the first group were performed in a location designated by the app within the city of Rio de Janeiro. We excluded users whose test results were inconclusive.

### Outcomes and variables

Our primary outcome was the test result (positive or negative) at the user level. Our goal was to identify clinical manifestations and individual factors associated with positive testing. Hence, we collected and assessed participant demographics (age, gender), nine symptoms (loss of smell or anosmia, fever, myalgia, cough, nausea, shortness of breath, diarrhea, coryza, and sore throat), and whether the user lives together with someone with a confirmed SARS-CoV-2 infection.

### Statistical analysis

We described the characteristics and symptoms of positive and negative tested participants, displaying the mean and standard deviation for continuous variables and the frequency for categorical variables. We then analyzed the individual association between symptoms and the test result using a logistic regression model adjusted to age and gender. We provided the corresponding Odds Ratio (OR) with a 95% confidence interval.

We aim to identify a combination of symptoms to build a prediction model for determining a participant with SARS-CoV-2 infection. For that, we compared five different machine learning techniques: Logistic Regression (stepwise), Naïve Bayes, Random Forest, Decision Tree (C 5.0), and eXtreme Gradient Boosting. To address the imbalanced response variable (only 11.8% are positive tests) during model training, we also evaluated four different data balancing techniques (downsampling, upsampling, SMOTE, and ROSE).

The dataset was randomly divided into training and test sets (ratio: 80:20). During model training, for each combination of machine learning and data balancing techniques, we applied grid-search hyperparameter optimization with 5-fold cross-validation, using the Area Under the ROC Curve (AUC) as the target metric. We comparatively evaluated the models’ performance in the test set and selected the one with the highest Matthews Correlation Coefficient (MCC)[5] value. The cut-off point for predicted values was 50% (i.e., participants with a probability higher than 50% were classified as “positive”, and “negative” otherwise). After our final model was incorporated by the “*Dados do Bem*” app, we performed an external validation using data from users in the state of *Goias*.

Finally, we evaluated the distribution of SARS-CoV-2 infection risks over the geographic area of the state of Rio de Janeiro modeled as a grid map (each grid is a 400m x 400m square area). Along with the participants with confirmed test results, we applied the chosen model to the sample of participants that had not been tested to obtain their estimated test result. We then calculated the proportion of estimated SARS-CoV-2 infections for each grid according to Equation 1.

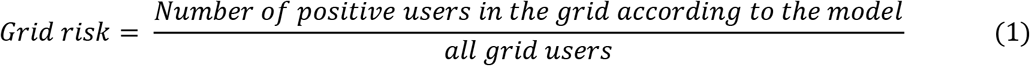

To avoid misinterpreting proportions in grids with scarce data, we considered grids with at least 10 participants (~94% of all observations). Then, we evaluated the distribution of the grid risks among all grids and classified them into five risk groups using the mean ± 0.5 and 1.5 standard deviations (SD) as thresholds: “very low” (< mean-1.5*SD), “low” (from mean-1.5*SD to mean-0.5*SD), “medium” (from mean-0.5*SD to mean+0.5*SD), “high” (from mean+0.5*SD to mean+1.5*SD), and “very high risk” (>mean+1.5*SD). Using this classification, we built a risk map for the state of Rio de Janeiro.

All analyses were performed in R 3.6.3, using ‘*tidyverse*’ package for data wrangling and plots; and ‘*caret*’ for the prediction models, with ‘*ranger*’ for Random Forest, ‘*C50*’ for Decision Trees, ‘*xgbTree’* for the eXtreme Gradient Boosting, and ‘*naivebayes’* for the Naïve Bayes model.

### Ethics Statement

The study is retrospective and had no human interference. All data acquired were anonymized, and the “*Dados do Bem*” app follows the Brazilian General Data Protection Regulation (*Lei Geral de Proteção de Dados* - LGPD). All users provided informed consent of de-identified data-use to non-commercial research upon registration in the app. All answers were optional.

### Data Availability

The data that support the findings of this study are available from “*Dados do Bem*” but restrictions apply to the availability of these data, which were used under license for the current study, and so are not publicly available. Data are however available from the authors upon reasonable request and with permission of “*Dados do Bem*”.

## Results

### Characteristics and self-reported symptoms associated with SARS-CoV-2 infection

From April 28, 2020, to July 16, 2020, 337,435 individuals registered their symptoms through the smartphone app. Of these, 49,721 users were already tested, from which 5,888 (11.8%) received a positive result for SARS-CoV-2 infection.

According to the self-reported information (Table 1), most participants were women (61.9%), health professionals (55.8%), with a median age of 41 years old (IQR: 33-51). Among those who tested positive for SARS-CoV-2 infection, cough was the most frequent symptom (59.6%), followed by myalgia (57.4%), coryza (56.3%), loss of smell/anosmia (52.9%), and fever (44.8%). When evaluating the association between each symptom and the test result, adjusted for age and gender (Fig 1), we found a similar result: loss of smell (odds ratio [OR]: 4.6; 95% CI: 4.4 - 4.9), fever (OR: 2.6; 95% CI: 2.5 - 2.8), and shortness of breath (OR: 2.1; 95% CI: 1.6 - 2.7) were associated with a positive result for SARS-CoV-2 infection.

**Table 1:** Characteristics and symptoms of the study population tested for SARS-CoV-2 infection. Results are displayed in median (interquartile range, IQR) for continuous variables and as percentage values for categorical variables.

|  | Total | Positive test | Negative test |
| --- | --- | --- | --- |
| **Participants n(%)** | 49,721 | 5,888 (11.8) | 43,833 (88.2) |
| **Characteristics** |  |  |  |
| Female, n(%) | 30,769 (61.9) | 3,641 (61.8) | 27,128 (61.9) |
| Age (years), median [IQR] | 41 [33-51] | 43 [34-53] | 40 [33-51] |
| Lives with a SARS-CoV-2 infected person, n(%) | 20,944 (42.1) | 3,398 (57.7) | 17,546 (40.0) |
| Health professional, n(%) | 27,737 (55.8) | 3,099 (52.6) | 24,638 (56.2) |
| **Self-reported symptoms, n(%)** |  |  |  |
| Coryza | 25,973 (52.2) | 3,315 (56.3) | 22,658 (51.7) |
| Cough | 23,430 (47.1) | 3,507 (59.6) | 19,923 (45.5) |
| Myalgia | 20,858 (42.0) | 3,380 (57.4) | 17,478 (39.9) |
| Sore throat | 20,794 (41.8) | 2,459 (41.8) | 18,335 (41.8) |
| Fever | 13,042 (26.2) | 2,640 (44.8) | 10,402 (23.7) |
| Diarrhea | 12,573 (25.3) | 1,778 (30.2) | 10,795 (24.6) |
| Loss of smell | 11,835 (23.8) | 3,112 (52.9) | 8,723 (19.9) |
| Nausea | 6,461 (13.0) | 1,025 (17.4) | 5,436 (12.4) |
| Shortness of breath | 354 (0.7) | 74 (1.3) | 280 (0.6) |
| No symptoms above | 10,865 (21.9) | 844 (14.3) | 10,021 (22.9) |

**Fig 1.**
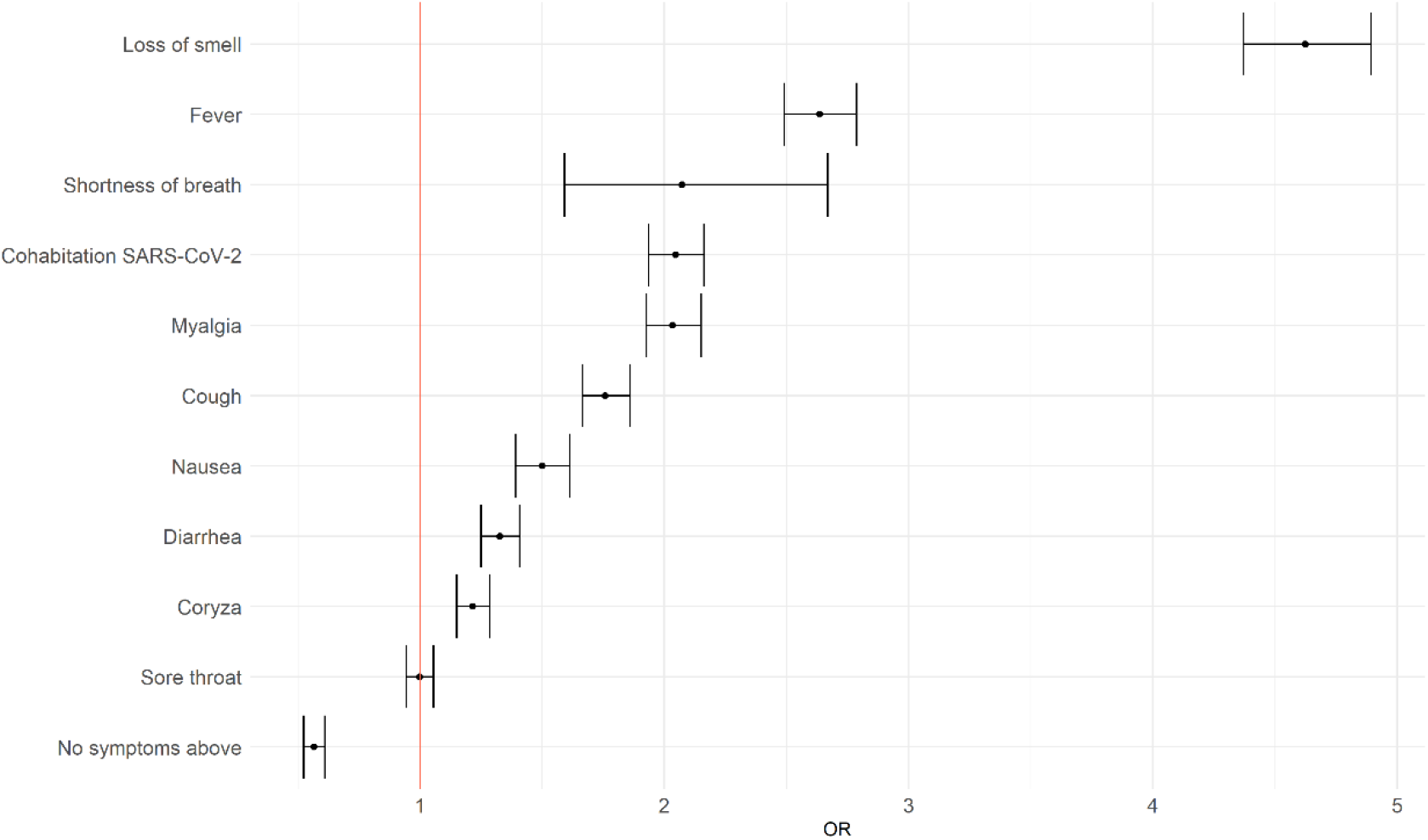
Association between symptoms and the SARS-CoV-2 infection. The Odds Ratio (OR) with 95% confidence intervals for each symptom using a logistic regression model adjusted to age and gender.

### Combination of symptoms and predictive modeling

Aiming to develop a model to predict positive participants based on the available dataset, we ran 25 different scenarios, combining machine learning techniques and sampling strategies. The logistic regression, gradient boosting and random forest techniques presented the best median MCCs (0.25), followed by the decision tree, and naïve Bayes (see the Supporting Information S2). Since the logistic regression model is more interpretable and parsimonious, we chose it as the final model (Equation 2).

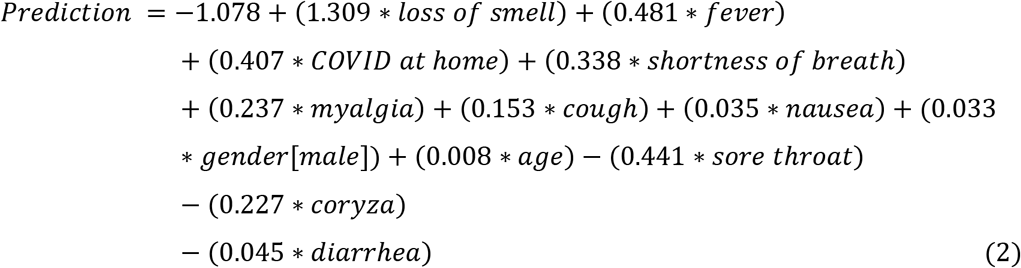

Regarding the classification metrics, our model performed as follows: sensitivity of 0.60; specificity of 0.75; PPV of 0.25; and NPV of 0.93. Thus, evaluating the predicted probability values, we observed that our model correctly predicts almost all negative tests with only 7% of false-negative users among the predicted as negatives (NPV = 0.93). We focus on avoiding false negatives (thus maximizing NPV) since we aim to prioritize tests in people with a potential positive result for COVID-19.

The characteristics of the false-negative and false-positive cases predicted by our model can be seen in the Supporting Information S3. We observed that most of the false-positive cases present the top-four predictors with the highest positive coefficients. At the same time, only four of the false negatives reported the loss of smell - the strongest predictor of a positive test.

### SARS-CoV-2 risk areas in Rio de Janeiro

Aiming to optimize the testing strategy, we applied the predictive model (Equation 1) to the 287,714 individuals registered in the app and still untested for SARS-CoV-2. According to our model, 99,431 (34,5%) of these participants were classified as a potential positive infection. We also visualized the geographical distribution of SARS-CoV-2 infection, calculating by Equation 2 the proportion of positive test results for each specific geographical area (named a grid) in the state of Rio de Janeiro. We calculated the predicted infection risk for each grid (Fig 2) and their risk groups. As of July 16, 2020, we observed that the southern areas in the city of Rio de Janeiro presented lower proportions of potential positive participants compared to the northern areas.

**Fig 2.**
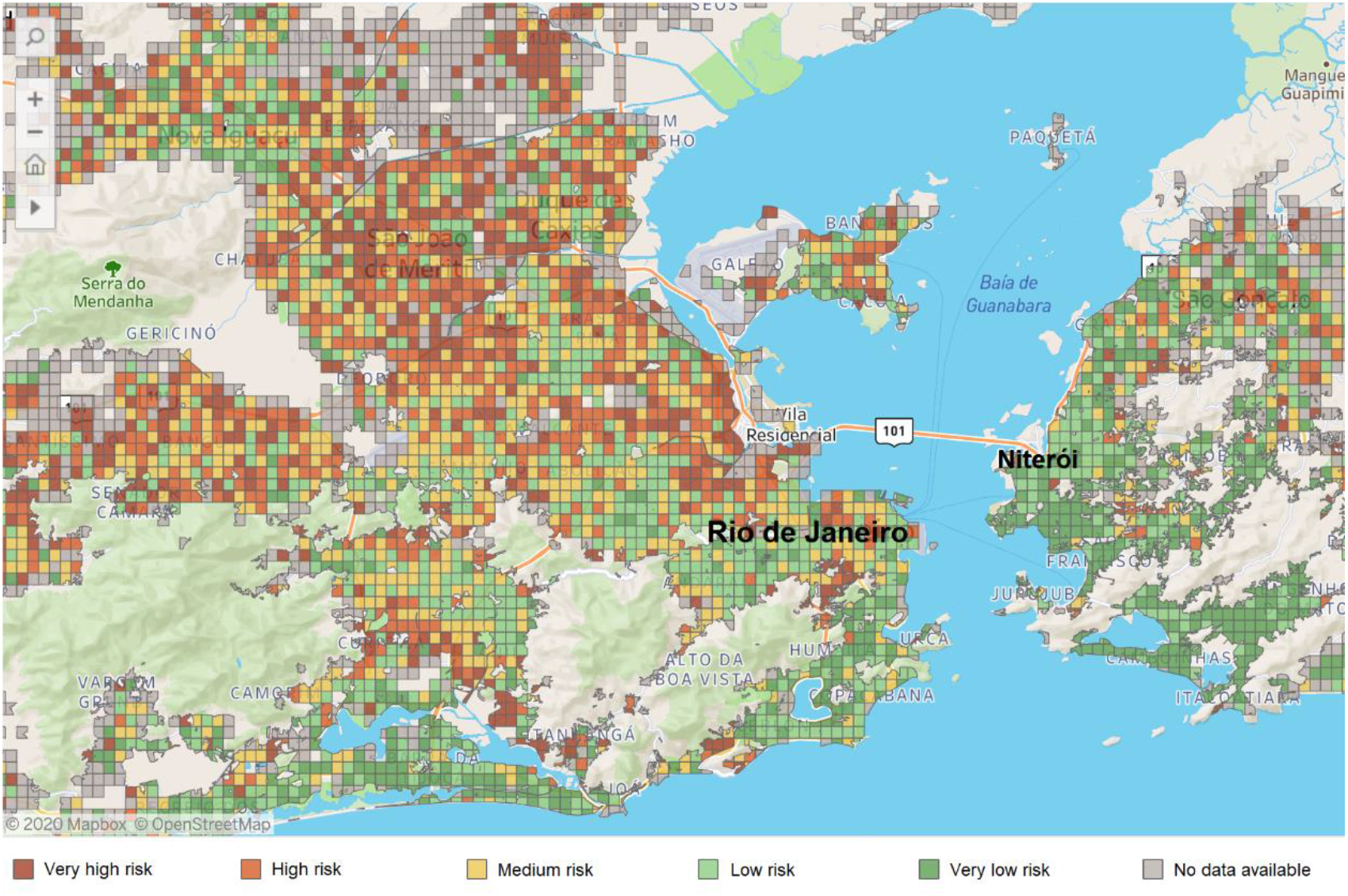
SARS-CoV-2 infection risk map (grid) of Rio de Janeiro state, displaying the city of Rio de Janeiro (capital, left) and the city of Niteroi (right) The grid risk considered the proportion of potential positive infection (observed test results + estimated from the prediction model) for each grid (400mx400m area). The risk groups were obtained as very low (<17%), low (≥17% and <33%), medium (≥33% and <48%), high (≥48 and <63%), and very high (≥63%). The map shows the distribution of risks as of June 10, 2020.

### External Validation

The “*Dados do Bem*” app incorporated our final model (Equation 2), using it to prioritize users for testing in some Brazilian states. For the external validation of the model, we analyzed the *Goias* state, where 465 registered users have been tested in 20 days. Of these, 52% had positive results.

## Discussion

Extensive testing programs for SARS-CoV-2 are, in general, not available in low- and middle-income countries, conferring the under-reporting of confirmed cases into a problem. A previous study estimated that only 9,2% of the Brazilian cases are being notified [6]. Restricting tests hinders the monitoring of the epidemic progression, resource planning, and evaluation of the effectiveness of the control measures. Besides, it leads to false conclusions that the disease is under control.

Since it is not possible to test all individuals, some studies suggest that the combination of symptoms could be used as a screening tool to identify people with potential SARS-CoV-2 infection who could be selected for testing [2,7]. This can be useful for planning public policies and for preventing the spread of the pandemic. That said, our study used data on individual symptoms and demographics obtained from an app-based system, known as “*Dados do Bem*”, to develop a model that predicts individuals with a higher probability of testing positive for SARS-CoV-2 infection.

Some works criticize the use of symptom-based screening strategies to quantify an individual likelihood of having COVID-19, due to the non-specific nature of some symptoms and the existence of co-infections with other respiratory viruses [8]. However, our results evidenced that such a strategy contributes to optimizing the overall testing strategy. Out of the 287,714 new users still not tested, our model estimated that the virus could infect 99,431 who, therefore, should be prioritized for testing. It reduced the need for extensive testing to only 34.5% of the registered untested users. This is undoubtedly beneficial as a public policy, especially in Brazil, a country with the second-highest number of COVID-19 cases and one of the lowest test rates^1^.

Our model was incorporated into the app and used to select patients for testing in the state of *Goias*. Of the first 465 users selected, 52% were tested positive. This positivity rate is extremely high compared to the observed positivity rate without a model (11.8%). It indicates that our model presented an excellent performance, improving the test strategy and selecting the users most likely to be positive in the actual scenario.

In addition to forecasting the likelihood of each user to acquire the virus, our model also assesses the geographical distribution of these participants, being a source of information to build a risk map for the state of Rio de Janeiro, as shown in Fig 2. The “*Dados do Bem*” app currently uses this map for categorizing risk areas, thus supporting decision-makers to identify areas with a higher risk of infection and accordingly refine testing and disease control strategies.

The risk map analysis developed in this work is exemplified in Fig 3, which presents the risk map of the south zone of the city of Rio de Janeiro. The chosen area includes both high-income neighborhoods (such as “*Ipanema,”* “*Leblon,”* and “*Gávea”*) and slums (such as “*Rocinha”*). The selected grid in Ipanema is classified as “low risk,” which means that the proportion of positive tests in this grid was between 17% and 33%. The other selected grid is in Rocinha, which, although located less than three miles from Ipanema, is classified as a “very high risk” grid, meaning that the proportion of positive tests living in this grid was higher than 63%. The existence of higher-risk areas in poor communities was also noted in other regions of the state (Fig 2). Many higher-risk grids were located in the north zone of the city, where the most deprived communities are located (“*Complexo do Alemão*” and “*Complexo do São Carlos,”* for example). The presence of social inequalities in Brazil has been pointed out by previous studies [9–11], which noted that it could be associated with the spreading of the disease.

**Fig 3.**
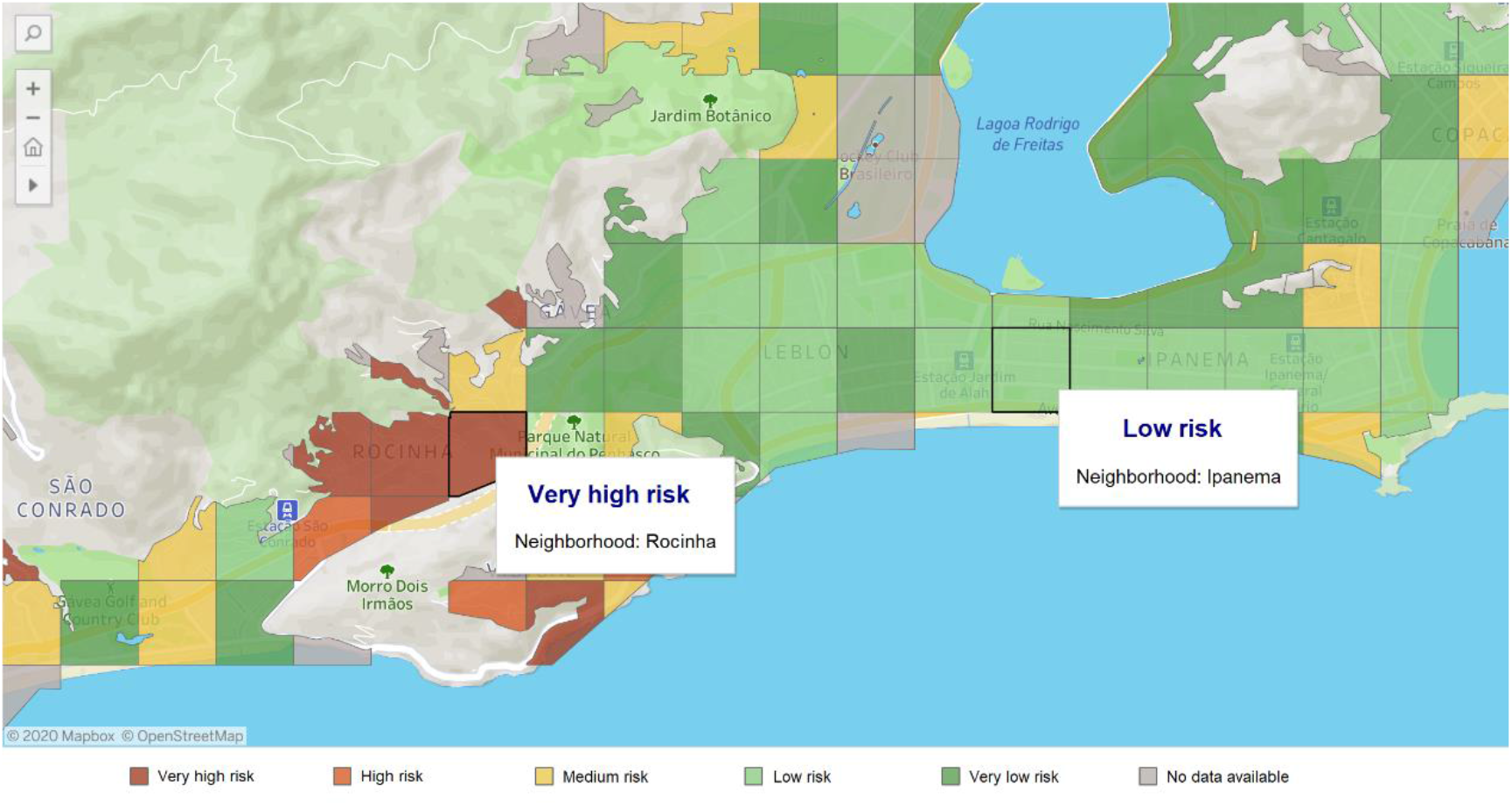
Risk map of two neighborhoods of Rio de Janeiro (Rocinha – very high risk and Ipanema – low risk)

Regarding our results of the reported symptoms, loss of smell (anosmia) was the strongest indicator of SARS-CoV-2, followed by fever, shortness of breath, myalgia, cough, nausea, diarrhea, and coryza. The significant influence of loss of smell and cough is in line with previous studies carried out in high-income countries such as the US and UK [2,12–14], and the influence of fever, myalgia, and nausea was pointed as significant in some studies [13,14], but others noted as not associated [2,12]. Regarding sore throat, diarrhea, and shortness of breath, previous studies observed that they are not significant predictors for the SARS-CoV-2 infection [2,12–14].

Menni and colleagues[2,7] used real-time tracking of self-reported symptoms similar to ours to predict potential SARS-CoV-2 infection in a cohort of individuals from the US and UK. This model was applied to compare the incidence in the UK regions. The authors noted that, in southern Wales, users reported symptoms that predicted, 5 to 7 days in advance, two spikes in the number of confirmed positive SARS-CoV-2 infection reported by public health authorities. The prediction models presented NPV of 0.75 and 0.87 in the UK (15,638 participants) and in the US (2,763 participants), respectively. Compared to them, our best model obtained a competitive performance (NPV of 0.93).

Sebo and colleagues [14] studied a sample of 1,543 primary care patients tested in two laboratories in the Lyon area (France) and found that the two symptoms most strongly associated with a positive test were loss of taste (ageusia) and loss of smell and that the combination of these symptoms resulted in an even stronger association (i.e., the odds of having a positive test were six times greater than the odds of having a negative test). A recent literature review of studies analyzing the presence of loss of taste and smell in SARS-CoV-2 infected patients concluded that, from a total of 10,818 patients, 8,823 presented ageusia (81.6%) and 8,088 presented anosmia (74.8%) [15]. Our results reinforce the literature conclusions about the strong influence of loss of smell.

This study presents some limitations. First, the symptoms are self-reported. Hence, the participant may report apparent manifestations of the disease, which may not be precise as physiological evaluation by a physician. Second, we were unable to know when a symptom appeared to indicate the stage of the disease at the testing moment. Third, a non-negligible number of false negatives may be present, considering the sensitivity of the serological test. However, identification of potential clusters and optimization of testing resources using a combination of self-reported symptoms is a viable strategy for many countries. A similar combination of symptoms can explain the SARS-CoV-2 infections in developed countries, such as the United Kingdom and the United States, as well as in LMIC such as Brazil.

## Conclusions

Our work used data regarding individual symptoms and demographics obtained from an app-based system to predict individuals with a higher probability of being infected by SARS-CoV-2. The model was applied in practice to select users from the state of Goias for testing, resulting in a positivity rate that was 4.4 more accurate than the positivity rate observed without using the model. In addition, the developed model was applied to new users of Rio de Janeiro to determine the risk of being infected in different geographical areas of this state. Our model estimated that 35.6% are potentially infected, reducing the need for extensive testing and optimizing the overall testing strategy. Based on the risk map derived from the model, the decision-makers may locate regions with a higher risk of positive tests, therefore allowing better testing and disease control policies.

## Data Availability

The data that support the findings of this study are available from "Dados do Bem" but restrictions apply to the availability of these data, which were used under license for the current study, and so are not publicly available. Data are however available from the authors upon reasonable request and with permission of "Dados do Bem".

## Acknowledgments

We would like to thank all collaborators from “Dados do Bem”, which specified and developed the application, and supported data collection. This work was supported by Conselho Nacional de Desenvolvimento Científico e Tecnológico (CNPq) (Grant numbers 169049/2017-5 to L.F.D., 306802/2015-5 and 403863/2016-3 to S.H., 310523/2016-8 to F.A.B., 144636/2019-0 to I.T.P.); Coordenação de Aperfeiçoamento de Pessoal de Nível Superior (CAPES) – Finance Code 001 (Grant number 88887.178844/2018-00 to L.S.L.B); Fundação de Amparo à Pesquisa do Estado do Rio de Janeiro (FAPERJ); and Pontifícia Universidade Católica do Rio de Janeiro (PUC-Rio).

## Supporting information

**S1 Fig. Pages of the COVID Symptom Tracker app**.

**S2 Table. Performance of the ML models computed from the independent test set for each combination**.

**S2 Fig. Box-plots are representing the MCCs of each strategy (points) for all methods**.

**S3 Table. Characteristics and symptoms of false-negative and false-positive users predicted from our model**.

**S3 Fig. Probability density function and frequency of the predicted values by the model using a testing set, compared to the real (observed) values**. The black vertical line corresponds to the cut-off of 0.5, and the colored dashed vertical lines correspond to the expected average probability for the group of negative (red) and positive (blue) groups.

## References

1. Coronavirus Update (Live). [cited 7 Jul 2020]. Available: https://www.worldometers.info/coronavirus/

2. Menni C Valdes AM Freidin MB Sudre CH Nguyen LH Drew DA et al. Real-time tracking of self-reported symptoms to predict potential COVID-19. Nature Medicine. 2020; 1–4. doi:10.1038/s41591-020-0916-2

3. Dados do Bem. Available: https://dadosdobem.com.br/

4. Acurácia dos testes diagnósticos registrados na ANVISA para a COVID-19. Available: https://portalarquivos2.saude.gov.br/images/pdf/2020/June/02/AcuraciaDiagnostico-COVID19-atualizacaoC.pdf

5. Chicco D Jurman G. The advantages of the Matthews correlation coefficient (MCC) over F1 score and accuracy in binary classification evaluation. BMC Genomics. 2020;21: 6. doi:10.1186/s12864-019-6413-7

6. Prado MF do, Antunes BB de P Bastos L dos SL Peres IT Silva A de AB da, Dantas LF et al. Analysis of COVID-19 under-reporting in Brazil. Revista Brasileira de Terapia Intensiva. 2020;32. doi:10.5935/0103-507X.20200030

7. Drew DA Nguyen LH Steves CJ Menni C Freydin M Varsavsky T et al. Rapid implementation of mobile technology for real-time epidemiology of COVID-19. Science. 2020;368: 1362–1367. doi:10.1126/science.abc0473

8. Callahan A Steinberg E Fries JA Gombar S Patel B Corbin CK et al. Estimating the efficacy of symptom-based screening for COVID-19. npj Digital Medicine. 2020;3: 1–3. doi:10.1038/s41746-020-0300-0

9. Lancet T. COVID-19 in Brazil: “So what?” The Lancet. 2020;395: 1461. doi:10.1016/S0140-6736(20)31095-3

10. Abrams EM Szefler SJ. COVID-19 and the impact of social determinants of health. The Lancet Respiratory Medicine. 2020;0. doi:10.1016/S2213-2600(20)30234-4

11. Ahmed F Ahmed N Pissarides C Stiglitz J. Why inequality could spread COVID-19. Lancet Public Health. 2020;5: e240. doi:10.1016/S2468-2667(20)30085-2

12. Brandstetter S Roth S Harner S Buntrock‐Döpke H Toncheva AA Borchers N et al. Symptoms and immunoglobulin development in hospital staff exposed to a SARS-CoV-2 outbreak. Pediatric Allergy and Immunology. n/a. doi:10.1111/pai.13278

13. Allen WE Altae-Tran H Briggs J Jin X McGee G Raghavan R et al. Population-scale Longitudinal Mapping of COVID-19 Symptoms, Behavior, and Testing Identifies Contributors to Continued Disease Spread in the United States. medRxiv. 2020; 2020.06.09.20126813. doi:10.1101/2020.06.09.20126813

14. Sebo P Tudrej B Lourdaux J Cuzin C Floquet M Haller DM et al. Clinical characteristics of SARS-CoV-2 patients: a French cross-sectional study in primary care. In Review; 2020 Jun. doi:10.21203/rs.3.rs-34635/v1

15. Passarelli PC Lopez MA Mastandrea Bonaviri GN Garcia-Godoy F D’Addona A. Taste and smell as chemosensory dysfunctions in COVID-19 infection. Am J Dent. 2020;33: 135–137.

